# The impact of chemosensory dysfunctions on weight loss

**DOI:** 10.1101/2021.11.06.21266003

**Authors:** Dimitrios Daskalou, Julien W. Hsieh, Marianne Hugentobler, Basile N. Landis

## Abstract

**Background:** The role of chemosensory senses (olfaction, taste, and trigeminal) is crucial, and their dysfunctions profoundly affect the quality of life, potentially impacting eating behaviors. However, it is unclear which chemosensory symptoms could lead to undernutrition. This study aims to investigate which findings in patients’ smell and taste workup are predictors of weight loss.

**Methods:** This is a retrospective study based on a validated questionnaire consecutively given to adult patients presented in smell and taste consultations during a 10-year period. Psychophysical tests were used to measure chemosensory function (Sniffin’ Sticks, taste powder, and Taste Strips tests).

**Results:** We included 554 patients (307 females) with a median age of 51 years (IQR 23). Among them, 76 (13.7%) reported involuntary weight loss due to chemosensory disorders occurred over periods ranging from 3 to 36 months. We found that the odds of losing weight were 2.1 times higher when patients reported changes in aroma perception (p-value=0.012; 95% CI 1.15 - 3.83). Parosmia, but not phantosmia nor smell loss, was a significant predictor of weight loss (OR 2.22; p-value=0.015; 95% CI 1.17 - 4.2). Furthermore, the duration of symptoms for more than two years was protective for weight loss (OR 0.44; p-value=0.014; 95% CI 0.23 - 0.85). Regarding putative etiologies, post-traumatic chemosensory dysfunction was also a significant predictor (OR 2.08; p-value=0.039; 95% CI 1.04 - 4.16). Concerning psychophysical tests, we found that the probability of a patient to present weight loss increased by 8% for every 1-unit reduction in Taste Strips score (p-value=0.006; 95% CI 0.87 - 0.98).

**Conclusion:** We recommend investigating weight loss in smell and taste consultations, especially when patients report changes in aroma perception, parosmia, duration of symptoms for less than two years, head injury, and when low Taste Strips score is measured.

## Introduction

The chemosensory senses (olfaction, taste, and trigeminal) are crucial for the quality of everyday life. The trigeminal system senses pain, temperature, and airflow in the nasal cavity. It perceives food and beverage texture in the oral cavity (1,2), where the basic tastes (salt, sweet, sour, bitter, and umami) are detected by the gustatory system (3). The olfactory system can discriminate between trillions of different odors (orthonasal pathway) and food aroma molecules streaming from the oral cavity to the nasopharynx, then backward to the roof of the nose where olfactory receptors are located. These receptors activation produces fine taste perception beyond the basic taste qualities (retronasal pathway) (4,5).

Chemosensory dysfunction is common, affecting approximately 20% of the general population over the age of 40 years (6). Patients with chemosensory dysfunction may experience one or more of the following symptoms: smell loss (complete or partial), parosmia (distorted odorant perception in odor source presence), phantosmia (smell perception without odor source), aroma loss or distortion (diminished or distorted retronasal perception of food molecules), taste loss (complete or partial loss of salty, bitter, sweet, sour perception), parageusia (distorted taste perception in stimulus presence) or phantogeusia (taste and oral sensation without stimulus). Subjective alteration of trigeminal sensation is rarely reported (7). These symptoms may be caused by different etiologies, including sinonasal, head trauma, upper respiratory tract infection, idiopathic, drugs, neurological disorders, or a multitude of other rarer causes (8). Testing these senses separately is often needed because self-ratings might be insufficient for localization and quantification of the deficit (9,10).

The role of these chemical senses is essential, and their dysfunctions profoundly affect the quality of life (11–14). One of the major impacts concerns eating behaviors. Food perception is a multisensory experience; thus, chemosensory dysfunction can alter both anticipation (orthonasal smell) and experience of consumption of food and beverage (retronasal smell and taste) (15). The relationship between chemosensory dysfunction and alteration of appetite and diet is well documented in the literature, with reported frequencies of 18 to 67% in these patients (8,14,16–22). As a result, changes in eating behaviors may lead to undernutrition and eventually to weight loss. Regarding potential predictors of weight loss, some studies highlight the increased risk of undernutrition in individuals suffering from distorted or diminished perception of smell or taste (8,14,16,20,22–24). However, the evidence is scarce, and it is still unclear which specific findings in smell and taste workup put patients at higher risk of weight loss. For example, aroma perception is a crucial component of food enjoyment potentially contributing to weight loss (25), but the association between aromas loss, measured by a retronasal olfactory test, and weight loss has never been tested. Furthermore, based on our clinical experience, we hypothesize that patients with taste dysfunction are more prone to weight loss.

Taking into account that chronic disease-related undernutrition increases morbidity and mortality with severe socioeconomic impacts (26), we feel that more research is needed to uncover specific findings of smell and taste workup that contribute to weight loss. This study aims to investigate the association between weight loss and self-reported chemosensory complaints, chemosensory test results, and putative etiologies.

## Materials and methods

### Study design

This is a retrospective study based on the analysis of an extended version of a questionnaire created by the German Working group for Taste and Smell Disorders. The aim of this questionnaire was to help the physician assess patient’s smell and taste complaints (27). This questionnaire was given to all patients presented in smell and taste consultations at Geneva University Hospitals (tertiary care facility) in Switzerland, between 2003-2012. The study was approved by the institutional ethics review board and was conducted according to the Declaration of Helsinki on Biomedical Research Involving Human Subjects (Institutional review board approval No: 13-161).

### Subjects

We asked from a consecutive series of 554 adult patients presented with chemosensory complaints to complete the questionnaire. Patients who reported weight loss due to reasons other than chemosensory dysfunction were excluded from the study.

### Outcome measures

#### Questionnaire

This is a two-part questionnaire completed by the patients (part I) and by the physician (part II). It is an extension of an existing questionnaire designed to help physicians conduct and address patients’ chemosensory complaints (27). In brief, the first part included detailed questions covering chief chemosensory complaints (smell, taste, aroma) and symptoms such as smell loss, parosmia, phantosmia, aroma loss, aroma distortion, taste loss, para- or phantogeusia, unusual oral sensation, and duration of symptoms. The questionnaire was then returned to the physician, who completed in the patient’s presence, information about involuntary self-reported weight loss due to the chemosensory problem. If the patients reported weight loss, its quantification was recorded in kg. The investigator also reported information about comorbidities, physical examination findings, chemosensory psychophysical test scores (orthonasal smell, retronasal smell, and taste), and putative etiology.

#### Orthonasal smell test

We performed the Sniffin’Sticks test (Burghart, Wedel, Germany), which comprises the olfactory threshold (T), discrimination (D), and identification (I) subtests (28). The TDI score was calculated as the sum of the results obtained from the three subsets. In the case of results for each nostril separately, we chose the best side’s score as the overall value.

#### Retronasal smell test

We assessed the retronasal olfactory function by applying the standardized “taste powder” tool. This tool uses food-related aromas in powder form applied at the posterior part of the tongue. The subjects then select from four descriptors the corresponding aroma, as described by Heilmann et al. (29). The retronasal score is calculated as the sum of the correct answers from ten trials.

#### Taste test

Taste evaluation was based on filter paper strips (“Taste Strips”, Burghart, Wedel, Germany) impregnated with four concentrations of the four basic taste qualities. We applied the test stimuli in random order with increasing concentrations (four concentrations for each of the four tastes) and on both sides of the anterior third of the extended tongue. A total “Taste strips” score was obtained for each participant by summing the correct answers (30,31).

### Statistical analysis

We used Pearson’s chi-square and Mann-Whitney test to uncover differences between patients with and without weight loss regarding age, gender, chief complaint, chemosensory symptoms, duration, putative etiologies, diagnosed depression, and chemosensory test results. When the expected frequencies were not fairly large, we used Fisher’s exact test instead of Pearson’s chi-square test. Post-hoc analysis for Pearson’s chi-square was performed using squared adjusted residuals (or z-square), which was transformed into p-values using a formula integrated into SPSS. To reject the null hypothesis, the p-value of 0.05 was divided by the number of associations to determine the adjusted p-value cut-off as described by Beasley et al. (32). We used univariable logistic regressions to verify these associations, and by consulting stepwise regression, we created two multivariable regression models. However, to have adequate statistical power for the model in Table 2, we could reasonably accommodate, at most, eight independent variables (putative etiologies accounted for four independent variables), and thus, we created a nested model by removing smell loss and duration variables. We ran a likelihood ratio test comparing the model with more predictors (full model, AIC = 395.09) with the model with fewer predictors (nested model, AIC = 395.29), and we found that the variables smell loss and duration do not give extra information to predict weight loss when all the other variables were taken into account (p-value = 0.12). All the assumptions for the logistic regression model were checked and satisfied. To conduct all analyses, we employed SPSS version 26 and an up-to-date version of R (version 1.2.5033, “https://www.r-project.org”). The statistical significance was defined as p < 0.05 (two-sided). Figures were created with Prism 8.0 and Adobe Illustrator.

Data of patients who did not answer, answered “I do not know”, gave uninterpretable answers (e.g., multiple choices selected when only one was authorized), or did not undergo chemosensory tests were considered as missing values. The remaining data regarding weight loss were n = 554, while data for 508 participants were free of missing values for all the variables assessed in the model of Table 2. Missing values were detected for patients who did not undergo the full TDI battery test (remaining = 512), retronasal test (remaining = 460), and Taste strips test (remaining = 198) and thus, data for 144 participants were free of missing values for the three chemosensory tests in the model of Table 3.

## Results

### Demographics and clinical findings in patients with and without weight loss

The median age in patients with (n = 76) and without (n = 478) weight loss was 52 (IQR = 22.5) and 50 (IQR = 23.75), respectively. Both groups had a proportional number of females. In these groups, we analyzed the association between weight loss and chief complaints (smell, aroma, taste), disease duration, chemosensory symptoms (smell, taste, and aroma quantitative and qualitative dysfunctions; burning mouth, xerostomia, sensation of oral foreign body), depression, putative etiologies, orthonasal, retronasal smell and taste testing. The significant associations were depicted in Figure 1 and detailed in Table 1. Regarding putative etiologies, post-traumatic etiology was associated with weight loss (squared adjusted residuals = 10, p-value = 0.002, adjusted p-value cut-off = 0.0063) (Figure 1c). As for the Taste strips score, although the median scores were close (18 vs. 20), the two groups presented different distribution (bi-modal vs. uni-modal, respectively), and this difference in medians was significant (p-value = 0.027) (Figure 1d). The median weight loss was 6 kg (IQR = 6) occurred over periods of 3 months (n = 15), 4-6 months (n = 9), 6-12 months (n = 25), 12-24 months (n = 11), and 36 months (n = 1). One patient lost 60 kg in a year after bariatric surgery and was excluded from the study.

**Table 1.** Population characteristics and comparisons.

| Variables | Weight loss |  | p-value |
| --- | --- | --- | --- |
|  | Yes | No |  |
| **Median age in years (IQR)** | 52 (22.5) | 50 (23.75) | 0.28 |
| **Gender (% female)** | 55.3 | 56.6 | 0.82 |
| **Chief complaint (%)** |  |  |  |
| Smell | 81.6 | 89.1 | 0.06 |
| Aroma | 72.4 | 55.8 | 0.008 |
| Taste | 42.1 | 29.1 | 0.024 |
| **Chemosensory symptoms (%)** |  |  |  |
| Parosmia | 28.9 | 15.7 | 0.006 |
| Phantosmia | 17.1 | 23.6 | 0.21 |
| Smell loss | 82.2 | 80.2 | 0.77 |
| Aroma loss | 65.8 | 69.2 | 0.55 |
| Aroma distortion | 74.3 | 64.1 | 0.1 |
| At least one basic taste altered | 34.2 | 30.8 | 0.55 |
| **Qualitative taste complaint and oral sensation (%)** |  |  |  |
| Burning | 11.8 | 12.3 | 0.9 |
| Bitter | 15.8 | 11.5 | 0.29 |
| Salty | 5.3 | 6.3 | 0.99 |
| Sour | 7.9 | 9 | 0.75 |
| Xerostomia | 39.5 | 28.7 | 0.06 |
| Foreign body | 7.9 | 7.7 | 0.99 |
| **Etiologies (%)** |  |  | 0.006 |
| Sinonasal | 8.9 | 22.5 | 0.019<sup>+</sup> |
| Post-traumatic | 41.1 | 21.8 | 0.002<sup>+</sup> |
| Post-infectious | 17.9 | 18.3 | 0.93<sup>+</sup> |
| Idiopathic | 32.1 | 37.4 | 0.45<sup>+</sup> |
| **Duration > 2 years (%)** | 17.1 | 32.4 | 0.01 |
| **Depression (%)** | 5.3 | 5.4 | 0.99 |
| **Median psychophysical test scores (IQR)** |  |  |  |
| Sniffin' Sticks TDI score | 19.12 (17.12) | 20 (17.5) | 0.53 |
| Taste strips score | 18 (15) | 20 (10) | 0.027 |
| Retronasal "powder tool" score | 5 (4) | 5 (4) | 0.88 |

IQR interquartile range, TDI threshold, discrimination and identification
<sup>+</sup> The adjusted level of significance is $p < 0.0063$ for Pearson's chi-square post-hoc analysis

**Figure 1.**
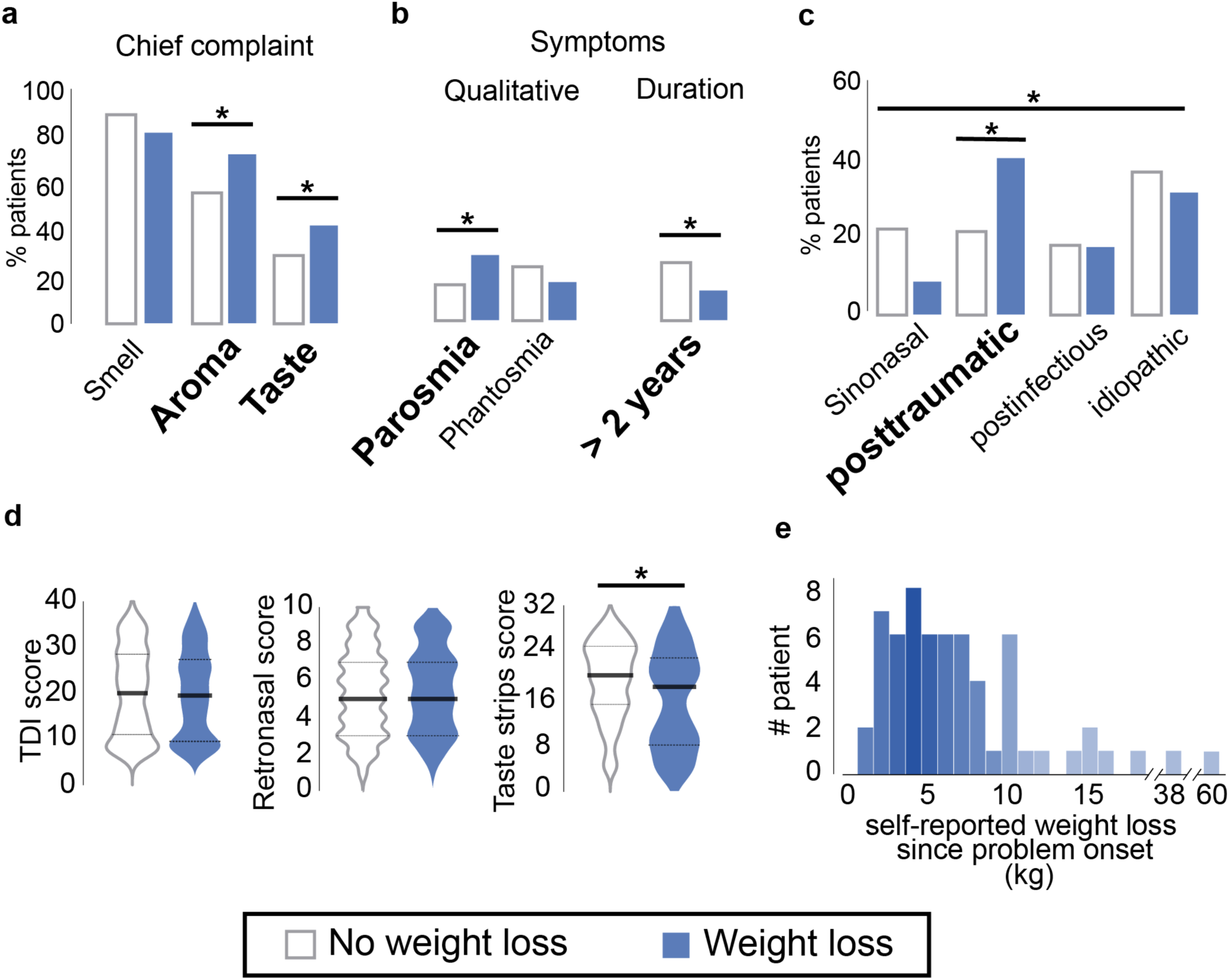
Findings of chemosensory workup between patients with (blue) and without (white) weight loss. (a) Comparison of relative frequencies between patients’ chief complaints and weight loss. (b) Association between qualitative olfactory dysfunctions (parosmia and phantosmia), duration of symptoms and weight loss. (c) Comparison between chemosensory dysfunctions putative etiologies and weight loss. (d) Presentation of chemosensory test scores with violin plots in patients with and without weight loss. (e) Absolute frequency of subjects presenting different degrees of weight loss in kg since the onset of the chemosensory complaint. Significant differences between the two groups are indicated by asterisks (* p-value < 0.05). TDI threshold, discrimination, and identification.

### Univariable analysis and multivariable logistic regression to predict weight loss

For chief complaint, we found that the odds to lose weight were 2.2 and 1.7 times higher for patients who reported alteration of aromas and taste, respectively (p-value = 0.006; 95% CI 1.25 - 3.88 and p-value = 0.043; 95% CI 1.02 - 2.86). Regarding the symptoms, the odds of presenting weight loss were two times higher for patients who presented parosmia (p-value = 0.013; 95% CI 1.17 - 3.63). Furthermore, duration of symptoms for more than two years was found to be protective for weight loss as the odds were 56% lower compared to patients with duration of symptoms less than two years (p-value = 0.014; 95% CI 0.23 - 0.85). Regarding putative etiologies, post-traumatic patients were at greater risk for developing weight loss (OR 2.17; p-value = 0.025; 95% CI 1.1 - 4.25) when compared with the reference group (idiopathic). We controlled for age as a possible predictor of weight loss, but the correlation was non-significant (p-value = 0.21; 95% CI 0.99 – 1.03). Thirty patients have been diagnosed with depression (four reported weight-loss), and we found no association between depression and weight loss (p-value = 0.99; 95% CI 0.32 – 3.05). After controlling for other variables, changes in aromas perception remained a significant predictor of weight loss (OR 2.1; p-value = 0.012; 95% CI 1.15 – 3.83). However, changes in taste perception proved non-significant in the multivariable model (p-value = 0.2; 95% CI 0.83 - 2.44). Regarding the chemosensory symptoms, self-reported parosmia continued to be a significant predictor (OR 2.22; p-value = 0.015; 95% CI 1.17 - 4.2), while phantosmia remained non-significant (p-value = 0.052; 95% CI 0.25 - 1.01). As for putative etiologies, the post-traumatic group presented a probability of losing weight two times higher than the reference group of idiopathic (p-value = 0.039; 1.04 - 4.16) (Table 2).

**Table 2.** Logistic regression models for weight loss prediction based on chief complaints, symptoms and putative etiologies (n = 508).

| Variables | Univariable analysis |  |  | Multivariable analysis |  |  |
| --- | --- | --- | --- | --- | --- | --- |
|  | OR | 95% CI | p-value | OR | 95% CI | p-value |
| **Smell loss** (yes / no) | 1.79 | 0.88, 3.62 | 0.11 |  |  |  |
| **Aromas perception complaint** (yes / no) | 2.2 | 1.25, 3.88 | 0.006 | 2.1 | 1.15, 3.83 | 0.012 |
| **Taste complaint** (yes / no) | 1.7 | 1.02, 2.86 | 0.043 | 1.42 | 0.83, 2.44 | 0.2 |
| **Parosmia** (yes / no) | 2.06 | 1.17, 3.63 | 0.013 | 2.22 | 1.17, 4.2 | 0.015 |
| **Phantosmia** (yes / no) | 0.67 | 0.35, 1.26 | 0.21 | 0.51 | 0.25, 1.01 | 0.052 |
| **Etiologies** (reference group = idiopathic) |  |  | 0.005 |  |  | 0.003 |
| Sinonasal | 0.45 | 0.16, 1.27 | 0.132 | 0.46 | 0.16, 1.32 | 0.15 |
| Post-traumatic | 2.17 | 1.1, 4.25 | 0.025 | 2.08 | 1.04, 4.16 | 0.039 |
| Post-infectious | 1.07 | 0.47, 2.44 | 0.868 | 0.76 | 0.32, 1.81 | 0.54 |
| Others | 1.96 | 0.93, 4.16 | 0.078 | 2.26 | 1.04, 4.91 | 0.04 |
| **Duration of symptoms**<br>(> 2 years / < 2years) | 0.44 | 0.23, 0.85 | 0.014 |  |  |  |
| **Age** (in years) | 1.01 | 0.99, 1.03 | 0.21 |  |  |  |
| **Depression** (yes / no) | 0.99 | 0.32, 3.05 | 0.99 |  |  |  |

OD odds ratio, CI confidence interval

We then examined TDI, retronasal, and Taste strips scores as predictors of weight loss in a multivariable model. A patient’s probability of presenting weight loss increased by 8% for every 1-unit reduction in Taste strips score adjusted for all the other variables of the model (p-value = 0.006; 95% CI 0.87 - 0.98). We did not find a significant association between weight loss and the other psychophysical test scores (Table 3).

**Table 3.** Logistic regression models for weight loss prediction based on psychophysical test scores (n = 144).

| Variables | Univariable analysis |  |  | Multivariable analysis |  |  |
| --- | --- | --- | --- | --- | --- | --- |
|  | OR | 95% CI | p-value | OR | 95% CI | p-value |
| **Sniffin' sticks TDI score** | 0.99 | 0.96, 1.02 | 0.41 | 1 | 0.96, 1.05 | 0.71 |
| **Retronasal "powder tool" score** | 1.01 | 0.89, 1.13 | 0.99 | 1.06 | 0.88, 1.27 | 0.48 |
| **Taste strips score** | 0.93 | 0.89, 0.98 | 0.006 | 0.92 | 0.87, 0.98 | 0.006 |

OD odds ratio, CI confidence interval, TDI threshold, discrimination and identification

## Discussion

Our main findings were that aroma issues, parosmia, duration of symptoms less than two years, low Taste strips scores, and post-traumatic etiology were associated with weight loss.

This study aimed to sort out findings in patients’ smell and taste workup that may point towards weight loss, which could increase morbidity and mortality with socioeconomic impacts. Evidence in the literature is scarce; results and methods are heterogeneous, leading to clinical uncertainty about how to pinpoint patients at risk of weight loss.

Although undernutrition has been mainly studied with orthonasal smell and taste tests, there is a lack of evidence in the literature about the link between aromas perception, measured by psychophysical tests, and weight loss. We found that aroma issues, as a chief complaint, were associated with weight loss even after controlling for confounders such as pure taste complaint. Patients had 2.1 times higher odds of losing weight if they had this chief complaint. However, this finding was not supported by retronasal smell testing results. A possible explanation is that the “taste powder” tool used in the study may not detect subtle aroma perception changes as it uses suprathreshold concentration. It is also worth noting that aromas, as perceived by the retronasal route, are generally mistaken with basic taste detected in the oral cavity as the general population is unfamiliar with this distinction (33). The question used was intended to name the principal complaint but might be too vague for the patients to understand the concept of aromas. However, further detailed questions about taste and aroma loss or distortion did not show differences between patients with and without weight loss. It remains possible that taste dysfunction, as measured by Taste strips, could explain this finding.

Regarding taste and its impact on weight loss, we included in the analysis other modalities contributing to flavor perception because these chemical senses interact closely. The hypothesis of mutual chemosensory weakening is based on the projection of the three chemical senses to the orbitofrontal cortex, which is considered as the secondary olfactory and gustatory cortex (34,35). Besides, Mazzola et al. found that the mid-dorsal insula, which plays an essential role in flavor perception, has a spatial overlap between olfactory, gustatory, and oral somatosensory representation (36). Migneault-Bouchard et al. also previously showed that the three chemical senses tend to decrease proportionally across different smell loss etiologies (37). By controlling for ortho- and retronasal smell, which contribute highly to flavor perception along with other senses (e.g., trigeminal, vision), we mostly present the effect of taste dysfunction in weight loss. Our results suggest a negative correlation between weight loss and Taste strips score. This contrasts with findings from Roos et al. who did not find a significant correlation between Taste strips score and body-mass index (BMI) (24); or De jong et al. who conducted a study in an elderly population and also found no association between taste score and energy intake or BMI (17). However, they used an unvalidated taste test composed of commercially available products, potentially triggering multiple aspects of the chemical senses. Besides, both studies had low sample size to detect subtle differences in taste function. In contrast, our study was powered to detect a smaller effect size, and we used a validated and more subtle test. In line with our clinical experience in daily practice, we bring new evidence that a low Taste strips score could be an independent risk factor for weight loss.

Regarding parosmia, patients suffering from it are at greater risk for developing weight loss, even after controlling for other chemosensory symptoms and putative etiologies. Mattes et al. highlighted the higher risk of dietary dissatisfaction and weight loss due to smell and taste distortions, compared to a complete or partial loss of these functions (20). They argued that distortions are more challenging to cope with, and patients begin to develop aversions to specific foods. Especially, patients who report parosmia have a more significant impact on their quality of life than individuals with simple loss of smell (13,38,39). They present, with higher rates, mild depression, and more prominent difficulties coping with their olfactory dysfunction (13). According to our results, depression is not a confounding factor between weight loss and parosmia. They also support the current literature that parosmia is a risk factor for weight loss. Olfaction is an essential component of food perception experience. People with smell dysfunction report reduced enjoyment of food in approximately 70% (11,19–21). The reduction in food enjoyment forces patients to change their eating behavior and develop strategies to enhance food perception. Common strategies are adding more spices, sweeteners, or salt and focusing on foods’ texture (12). These strategies may be efficient for individuals with simple olfactory loss, to alter their diet, and to prevent caloric deficit and undernutrition. In addition, as our results suggest, disease duration of more than two years leads to a lower risk of weight loss. The importance of chemosensory senses in everyday life may decrease as a result of a “response shift”, which is defined by a recalibration of internal standards and values over time to incorporate a loss of function after neurological injury (40). Our data agree with the majority of previous studies (8,17,19,20,22). Neither self-reported smell loss nor low TDI score is linked with weight loss. Aschenbrenner et al. speculated that decreased or absent olfactory function could result in decreased bodyweight (16). However, patients with isolated congenital anosmia have no weight differences with aged-matched controls (41). Roos et al. measured TDI scores in patients with Parkinson’s disease and found a positive correlation between TDI score and BMI (24). However, the authors controlled only for disease duration, but not energy expenditure, which has been shown to increase during the course of the disease and is probably the main reason for weight loss (42).

Post-traumatic patients were at higher risk of losing weight when compared to other putative etiologies regardless of the impact of chemosensory symptoms. Interestingly, Crenn and colleagues showed in a longitudinal study that about 30% of patients with severe head trauma lose weight. They argue that this may be due to the disruption of pathways regulating food intake (43). In our study, 20% of patients lost weight among patients suffering from post-traumatic chemosensory loss, which is a finding that also seems to be independent of other chemosensory symptoms.

Our study has several limitations. First of all, as a retrospective study has known inherent biases. We took steps to minimize selection bias by including consecutive patients. Data were also carefully and regularly reported in the study database. However, weight loss was self-reported in a single consultation and not measured and documented in follow-up visits. Although several confounders were tested and included in our regression models, we cannot exclude others that may influence our conclusions, such as trigeminal dysfunction that was not measured. Also, we have to interpret the external validity of our analysis critically. Although one in five people present chemosensory disorders in the general population, only a few are presented in specialized consultations. We can argue that these patients are motivated because of the higher impact of their complaints in everyday life. As a result, conclusions shall be drawn for patients seeking medical help for their chemosensory complaints and not for the general population.

## Conclusion

We highlighted the different findings in chemosensory disorders workup that point towards weight loss. We recommend investigating weight loss in smell and taste consultations, especially when patients report changes in aroma perception, parosmia, duration of symptoms for less than two years, head injury, and when low Taste strips score is measured.

## Data Availability

All data produced in the present study are available upon reasonable request to the authors

